# Sex specific trajectories of verbal memory decline suggest masking of Alzheimer’s Disease symptoms in females

**DOI:** 10.1101/2025.11.07.25339777

**Authors:** Sasha Novozhilova, Vladimir Fonov, Neda Shafie, Denise Klein, Sylvia Villeneuve, D. Louis Collins, the ADNI and PREVENT-AD research groups

## Abstract

**INTRODUCTION:** Verbal memory tests like the Rey’s Auditory Learning Test (RAVLT) are widely used in Alzheimer’s disease assessments without consideration of females’ well-known advantages in verbal memory. This study explores if this form of cognitive reserve masks symptoms and delays diagnosis in females.

**METHODS:** We employed a retrospective longitudinal cohort design (ADNI, PREVENT-AD) to model Immediate Recall subcomponents on the RAVLT over time in amyloid negative (Aβ-) cognitively normal participants versus Aβ+ participants that progressed to AD dementia.

**RESULTS:** Our Bayesian model successfully identified the inflection point where cognition deviates from normal onto the AD dementia trajectory. The timing of decline onset differed significantly across sexes, with Aβ+ females maintaining normal cognition longer than Aβ+ males. Among individuals with AD dementia, females experienced a steeper annual decline than males.

**DISCUSSION:** Delayed onset of decline suggests that females preserve cognition despite neuropathology accumulation. This masked period represents a window for earlier detection and timely intervention before irreversible neurodegeneration.

**Research in Context:**

1. Systematic review: The authors reviewed the literature from PubMed and Elsevier to identify verbal memory as a core feature in the Alzheimer’s Disease discourse. Our critical appraisal was complemented with meetings with expert neuropsychologists and neuroinformaticians.
2. Interpretations: We find that females have superior performance on Rey’s Auditory Learning Test (RAVLT) Immediate Recall, females maintain performance for longer than males before being diagnosed with Alzheimer’s Disease, and once diagnosed, they decline faster than males. These findings underscore the importance of conducting Alzheimer’s Disease assessments with considerations for sex differences.
3. Future directions: We aim to continue the investigation of cognitive reserve in females in the context of the Amyloid, Tau and Neurodegeneration (ANT) framework. Considering the early stage of disease is of interest given that our results show the appearance of cognitive symptoms many years before official dementia diagnosis.

**Highlights:**

- In cognitively healthy amyloid negative participants, females outperform males on the Rey’s Auditory Learning Test (RAVLT).
- In the amyloid positive participants, females maintain normal verbal memory for 2.7 years longer than males.
- After divergence from cognitive normality, females experience approximately a 25-50% steeper cognitive decline compared to males.
- Our findings call attention to the female verbal memory advantage as an influential form of cognitive reserve that requires diagnostic consideration.

## Background

In Alzheimer’s Disease (AD), cognitive reserve may reflect compensatory strategies that mitigate pathology’s cognitive impact. Traditional views of cognitive reserve focus on static factors such as occupational attainment or education that provide a baseline boost in cognition despite equal pathology (Boots et al., 2015; Stern, 2006, 2009). Cognitive reserve theory focuses on the dynamic nonlinear decline where individuals with high cognitive reserve have a delayed onset of cognitive symptoms followed by an accelerated loss of function (Emrani & Sundermann, 2025; Stern, 2009). Interestingly, female AD dementia patients have been described to suffer from more precipitous cognitive decline than men, possibly reflecting diagnosis at a later disease stage that may coincide with depletion of cognitive reserve (Lin et al., 2015; Nebel et al., 2018; Rosende-Roca et al., 2025). Working memory subprocesses may compensate for episodic memory loss in AD through executive control strategies (i.e. auditory attention span, interference control) that maintain functional performance despite accumulating pathology <u>(Kljajevic et al. 2023)</u>. This strategic form of cognitive reserve may explain why working memory has gained popularity as a prognostic tool to better predict global cognitive decline (Pillai et al., 2014).

Delayed diagnosis of female AD patients is of particular concern given the reliance of assessments based on verbal memory. The Rey’s Auditory Learning Test (RAVLT) is a particularly strong tool for measuring verbal cognition, encapsulating both episodic and working memory. However, the sex blind RAVLT fails to properly flag female cognitive decline and overestimates male cognitive impairment by 10% (Sundermann et al., 2019). Indeed, the literature points to females’ superior abilities in verbal memory as a form of sex-specific cognitive reserve that masks early signs of AD dementia (Emrani & Sundermann, 2025; Ferretti et al., 2018; Stricker et al., 2021; Sundermann et al., 2016). A series of cross-sectional studies have clearly demonstrated that females preserve superior verbal memory abilities at the amnestic mild cognitive impairment stage despite similar pathological burden compared to males; and that this advantage disappears at the onset of AD dementia (Banks et al., 2021; Caldwell et al., 2017; Gale et al., 2007; Sundermann et al., 2015, 2016, 2017, 2020). Furthermore, a longitudinal study of normal cognitive aging found sex to describe 8-10% of RAVLT performance variability (Stricker et al., 2021). Understanding the longitudinal sex-specific trajectories of verbal memory in AD is critical to identify cognitive vulnerabilities and precise intervention windows.

The primary aim of the current study is to characterize sex-specific critical periods in the transition from preclinical to clinical AD. We model two cognitive trajectories: unimpaired cognition in amyloid negative (Aβ-) subjects and progressive AD-related cognitive decline in Aβ+ subjects, to identify the temporal inflection point where the pathways diverge. We determine if there are sex differences for the timing of this inflection point or in the rate of cognitive decline afterwards. Our main hypotheses are that (i) cognitively unimpaired Aβ-females will demonstrate a longitudinal RAVLT advantage over males, establishing reserve differential; (ii) the timing of RAVLT decline will differ between Aβ+ males and females, with females maintaining performance for longer and thus experiencing delayed detection of clinically significant decline due to symptom masking; and (iii) following the inflection point, the slope of RAVLT decline will be sharper in Aβ+ females, reflecting rapid depletion of verbal ability and cognitive reserve. As a secondary aim, we investigate the cognitive processes that drive cognitive reserve by fractioning RAVLT scores. We hypothesize that sex differences in verbal memory trajectories will be most pronounced in Early Learning as it reflects working memory subprocesses.

## Methods

### Data Source

This paper utilized data obtained from the Alzheimer’s Disease Neuroimaging Initiative (ADNI) database (adni.loni.usc.edu). ADNI includes serial magnetic resonance imaging (MRI), positron emission tomography (PET) and other biological markers. The ADNI also makes clinical and neuropsychological assessments used to measure the progression of mild cognitive impairment (MCI) and early AD dementia openly available. ADNI was launched in 2003 as a public-private partnership and performs routine follow-ups on its participants. The study received ethical approval from the review boards of all participating institutions. Written informed consent was obtained from participants or their study partners. Participants were selected from ADNI-1, ADNI-GO, ADNI-2, and ADNI-3. Demographic, diagnostic, and APOE4 ε4 information was extracted from the ADNIMERGE table. RAVLT performance was obtained from the NEUROBAT table, and the CDR global score from the CDR table. PET Aβ status was obtained from UCBERKELEY_AMY_6MM table and Aβ CSF status was obtained from UPENNBIOMK9 for those who did not have PET data available. Participants in ADNI attended an average of 3.45 ± 1.63 follow-up visits.

To complement the cognitively unimpaired ADNI cohort, we included data from the PResymptomatic EValuation of Experimental or Novel Treatments for Alzheimer’s disease (PREVENT-AD) study (Villeneuve et al., 2025). The PREVENT-AD database includes neuroimaging, biological, and neuropsychiatric assessments from cognitively unimpaired individuals with a family history of sporadic AD dementia. The study received ethical approval from the McGill University review board, and all participants provided written informed consent. Data was obtained from the internal sharing repository (Villeneuve et al., 2025). PREVENT-AD participants attended an average of 3.45 ± 1.63 follow-up visits.

### Groups

To estimate the deviation from the normal trajectory of verbal memory in AD patients, we defined two groups based on amyloid status (Jack et al., 2024). The first group (n = 389, males = 182) is composed of cognitively unimpaired subjects with negative amyloid status (Aβ-, i.e. those without evidence of Alzheimer’s disease as determined by either PET or CSF) at all visits from ADNI and PREVENT-AD. Aβ status was assessed across all visits in both cohorts using PET or CSF. In ADNI AV45 (cut-off 1.11 SUVR), FBB (cut-off 1.08 SUVR) and NAV (cut-off 1.14 SUVR) Aβ PET tracers using the whole cerebellum as reference were used to binarize positivity thresholds. Additionally, to cover as many time points as possible we used a ratio of CSF pTau/Aβ(1–42; cutoff 0.028) when PET data was not available (Leuzy et al., 2023). We applied the study specific SUVR threshold to the PREVENT-AD NAV PET tracer (cut-off 1.26 SUVR) scans to define the Aβ status (Qiu et al., 2025; Yakoub et al., 2025). To simplify interpretation, in Table 1 we report Aβ volumes at baseline in Centiloids (Klunk et al., 2015). The second group (n = 492, males = 249) includes only amyloid positive (Aβ+) subjects from ADNI (i.e., those with evidence of Alzheimer’s disease) that either met neuropsychological criteria for dementia at baseline or at some point during follow-up (Jack et al., 2024). The onset of Alzheimer’s disease dementia was set based on the ADNI diagnostic criteria: when an Aβ+ participant manifested significant memory decline on the Wechsler Memory Scale (based on age and education) and abnormal global cognition measured by the Mini Mental Status Examination (MMSE<26) or Clinical Dementia Rating (CDR>0).

**Table 1.** Demographic Information. ***Demographic and clinical characteristics of the sample at baseline,*** stratified by sex, including age, education, cognitive scores, and APOE ε4 carrier status. Aβ values at baseline are reported in Centiloids and as a ratio of pTau/Aβ.

| | Group Aβ- (Controls) | | Group Aβ+ (AD Trajectory) | |
| --- | --- | --- | --- | --- |
| Variable | Female | Male | Female | Male |
| Participants, n | 280 | 212 | 191 | 249 |
| Age (mean ± SD) | 70.4 ± 6.2 | 72.3 ± 6.4 | 72.4 ± 7.4 | 74.8 ± 7.1 |
| Education (mean ± SD) | 15.9 ± 2.9 | 16.9 ± 2.8 | 15.2 ± 2.5 | 16.3 ± 2.8 |
| CDRSB | 0.0 ± 0.1 | 0.0 ± 0.1 | 2.7 ± 2.0 | 2.7 ± 1.8 |
| APOE ε4: 0 allele | 215 (76.4%) | 165 (77.8%) | 56 (29.3%) | 75 (30.1%) |
| APOE ε4: 1 allele | 62 (22.1%) | 45 (21.2%) | 104 (54.5%) | 121 (48.6%) |
| APOE ε4: 2 alleles | 4 (1.4%) | 2 (0.9%) | 31 (16.2%) | 53 (21.3%) |
| PET Centiloids (CL) | 6.9 ± 12.0 | 3.8 ± 13.0 | 86.7 ± 33.8 | 85.9 ± 36.5 |
| CSF pTAU/ABETA | 0.016 ± 0.005 | 0.016 ± 0.009 | 0.068 ± 0.036 | 0.066 ± 0.022 |

### Statistical Analysis

Statistical modelling was performed using the NumPYRO package (version 0.19.0, Python version 3.12.6). Verbal memory was summarized using three outcomes reflecting different stages of cognitive processing. The immediate RAVLT recall trials were fractioned into Early Learning (average of trials 1 and 2), Late Learning (average of trials 4 and 5) and Total Learning (average of trials 1-5). In line with previous work (Almkvist et al., 2025), Early Learning was defined as indexing auditory-verbal attention and initial encoding capacity, which are often supported by executive strategies (e.g. active rehearsal, semantic organization). Late Learning, in contrast, reflects encoding efficiency and the transition to memory consolidation. Total Learning provides a global measure of the cumulative learning curve.

Each outcome was modeled using a hierarchical Bayesian model with repeated measures nested within participants (Eq. 1). In this framework, expected RAVLT performance for individual *i* was expressed as the sum of a sex-specific baseline, controlling for education and APOE4 ε4 genotype, a disease-related change component evolving over time relative to diagnosis of Aβ+ Dementia, plus residual error.

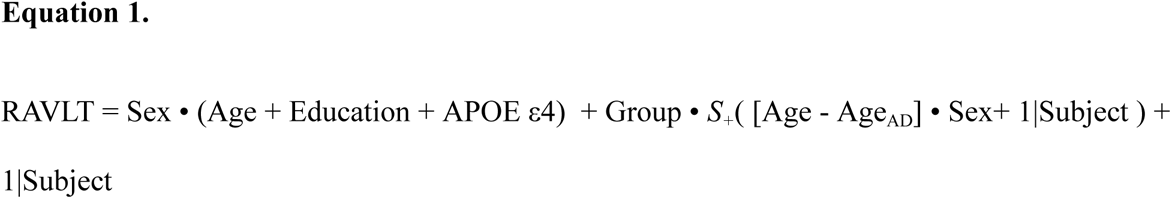

Where *Age* is the subject’s age at each visit, Group = 0 for Aβ- and =1 for Aβ+, so that SoftPlus function is applied only to the Aβ+ group, *S_+_* is the Softplus function, *Age_AD_*is the age at which they meet neuropsychological criteria for AD dementia. The SoftPlus function is continuous and differentiable, increasing monotonically from zero, thereby allowing both the timing and slope of decline to be estimated directly from the data (see Appendix for full specification). This disease-related term captures the gradual emergence of cognitive decline, avoiding abrupt changes at the point of diagnosis. The sharpness parameter controlling the width of the nonlinear transition was fixed at 10 years based on empirical evaluation.

All continuous predictors were standardized using z-scores derived from the mean and standard deviations of the Aβ- group at baseline. Posterior distributions were estimated using Markov chain Monte Carlo sampling. Effects were considered supported by the data when the 95% highest density interval (HDI) excluded zero. Model fit was assessed using the leave-one-out cross validation (LOO), with Pareto-k diagnostics used to identify influential observations. These diagnostics indicated that the models provided an adequate fit to the observed data (Vehtari et al., 2017).

## Results

### General model estimates

We report the overall model estimates, including baseline Total Learning performance and the effects of APOE ε4 genotype, education, age in Table 2. The effects of sex are reported in the section below. For visualization purposes, we include a zero-centered forest plot of the posterior distribution of the predictors in the Appendix (Fig. A1). Higher education was found to be positively associated with scores on RAVLT Total (β= 1.710, 95% HDI = [−1.935, −1.700]) and Late Learning (β= 0.359, 95% HDI = [−0.471, −0.416]) at baseline. Education was not found to be associated with Early Learning scores (β= 0.30, 95% HDI = [0.180, 0.416]) when estimating the effects for both males and females together. In both groups, APOE ε4 allele status did not have a significant effect on RAVLT performance. We identified a main effect of age negatively influencing RAVLT Total, Early and Late Learning scores in both the Aβ- and Aβ+ groups (Table 2).

**Table 2:** General Model Estimates. ***Main effects coefficients of predictors of RAVLT subscores.*** Here the values represent the posterior mean with 95% High Density Intervals (HDI) in brackets. The model was fit using MCM sampling and fit was assessed with Leave-One-Out (LOO) validation. Note. Main effects were estimated by averaging sex-stratified Numpyro model output

| Variables | Total Learning | Early Learning | Late Learning |
| --- | --- | --- | --- |
| **Intercept** | 45.830 [45.280<br>46.354] | 6.810 [6.705<br>6.908] | 11.218 [11.094<br>11.342] |
| **Aβ- Age (slope)** | -0.533 [-0.915<br>-0.106] | -0.00 [-0.051<br>0.053] | -0.231 [-0.321<br>-0.140] |
| **Aβ+ Age (slope)** | -1.816 [1.064<br>2.374] | -0.264 [-0.286<br>-0.241] | -0.443 [0.209<br>0.519] |
| **Education** | 1.710 [-1.935<br>-1.700] | 0.303 [0.180<br>0.416] | 0.359 [-0.471<br>-0.416] |
| **APOE ε4: 1 allele** | 0.012 [-0.838,<br>0.861] | 0.049 [-0.083,<br>0.286] | -0.031 [-0.250,<br>0.153] |
| **APOE ε4: 2 alleles** | 0.020 [-1.121<br>1.254] | 0.025 [-0.191,<br>0.272] | -0.018 [-0.327,<br>0.256] |

### Sex-stratified model estimates

Table 3 shows the sex-stratified coefficients for the models of each RAVLT outcome subscore. On Figure 1, we display the full fitted model, for Total, Early and Late learning. The slopes for normal control Aβ- group are represented as a dotted line with males in blue and females in pink; the slopes of the AD dementia Aβ+ group are drawn with solid lines in each sex’ respective color. For additional visualization, Figure 2 in the Appendix (Fig. A2) depicts a z-scored forest plot of the posterior distributions of the sex-stratified predictors. In the Aβ- group, the male-female differences in the Total Learning intercept of –4.73 (95% HDI = [−6.05, −3.38], Fig. 2) indicates that females outperformed males on average. This pattern persisted in Early (female-male difference = −0.77, 95% HDI = [−1.01, −0.53], Fig. A.1b) and Late Learning (male-female difference = −1.08, 95% [−1.39, −0.78], Fig. Fig. 2). In the Aβ- groups, the effect of age did not reach significance in Total (female-male difference = −0.07, 95% HDI = [−0.72, 0.60], Fig. A.2a), Early (male-female difference = −0.01, 95% HDI = [−0.12, 0.09], Fig. 2) and Late Learning (male-female difference = 0.02, 95% HDI = [−0.15, 0.19], Fig. 2). Sex differences in the rate of decline in the Aβ+ group are reported in the section below. After adjusting for education as a covariate, the analysis revealed no significant influence of education on Total (male-female difference = −0.89, 95% HDI = [−2.12, 0.32], Fig. 2) and Late Learning (female-male difference = −0.08, 95% HDI = [−0.35, 0.21], Fig. 2). The effect of education did reach marginal significance in Early Learning (male-female difference = −0.26, 95% HDI = [−0.48, −0.04], Fig. 2). We also showed that APOE ε4 allele status did not have a significant effect on the rate of decline. The male-female difference on Total Learning performance in carriers of one allele in slope was 0.19 (95% HDI = [−1.3, 2.1]). In subjects with two alleles the male-female in slope was 0.66 (95% HDI = [−1.3, 3.8]). Similar patterns of APOE ε4 allele status were observed for Early and Late Learning (Table 3).

**Figure 1.**
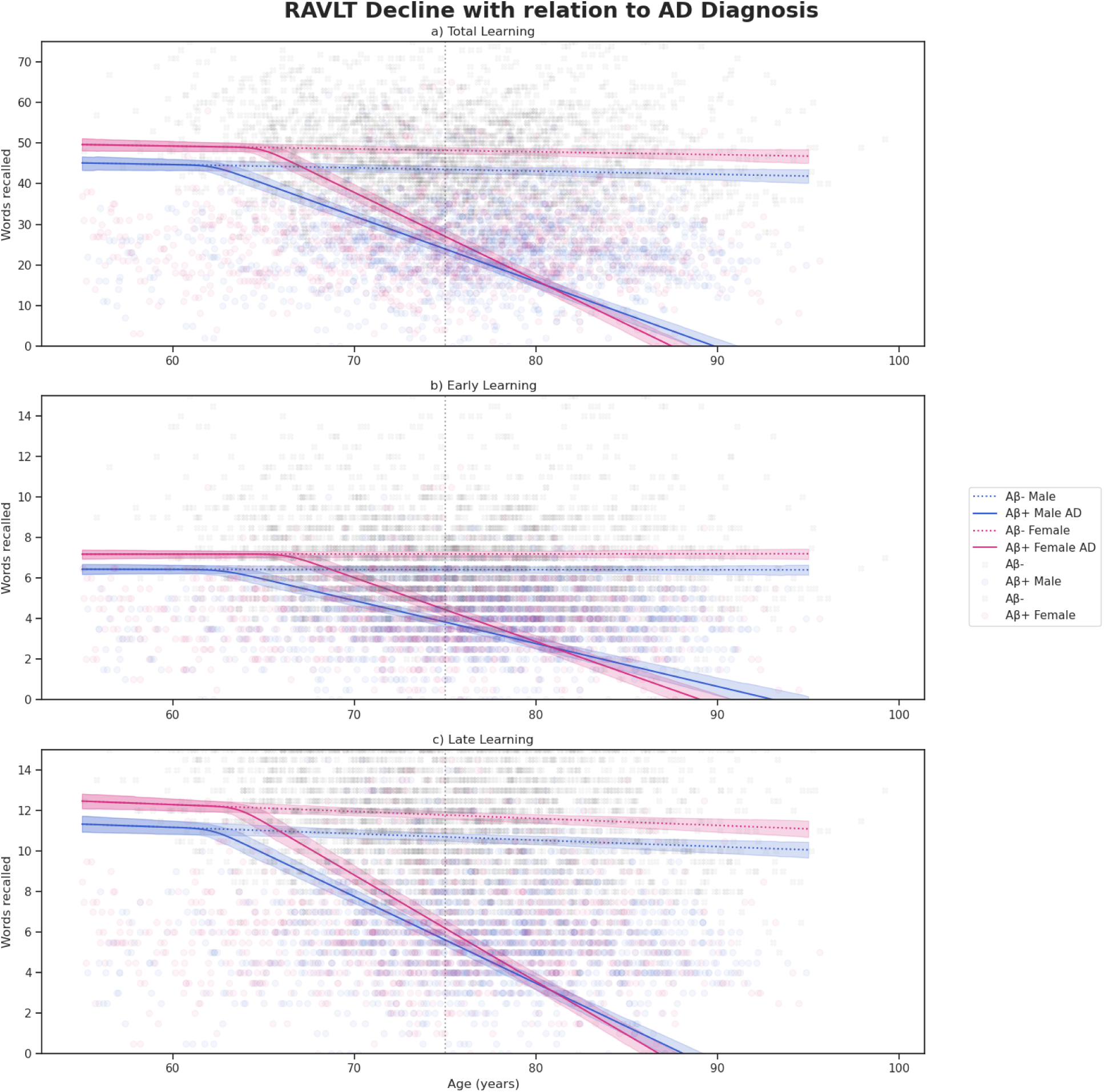
Posterior Predictive Trajectories of RAVLT Immediate Recall Performance Across Age by Sex and AD Progression for a given AD diagnosis at 75y. All models include Age, Education, and APOE ε4 as fixed effect covariates. Panel A shows Total Learning (sum of RAVLT trials 1-5), Panel B shows Early Learning (average of trials 1 and 2), and Panel C shows Late Learning (average of trials 4 and 5). Solid lines represent predicted mean trajectories for Aβ+ participants that progress to AD dementia, while dashed lines represent cognitively normal Aβ- participants. Blue lines correspond to males, pink lines to females. Shaded regions indicate 95% highest density intervals (HDI) of the posterior predictions. Raw data points are overlaid gray X-symbols for CN- and semi-transparent circles for Aβ+ participants in their respective color based on sex. Note. The dotted black line vertical at 75 years corresponds to a counter-factual onset of AD dementia, since diagnosis time is variable for each subject.

**Figure 2.**
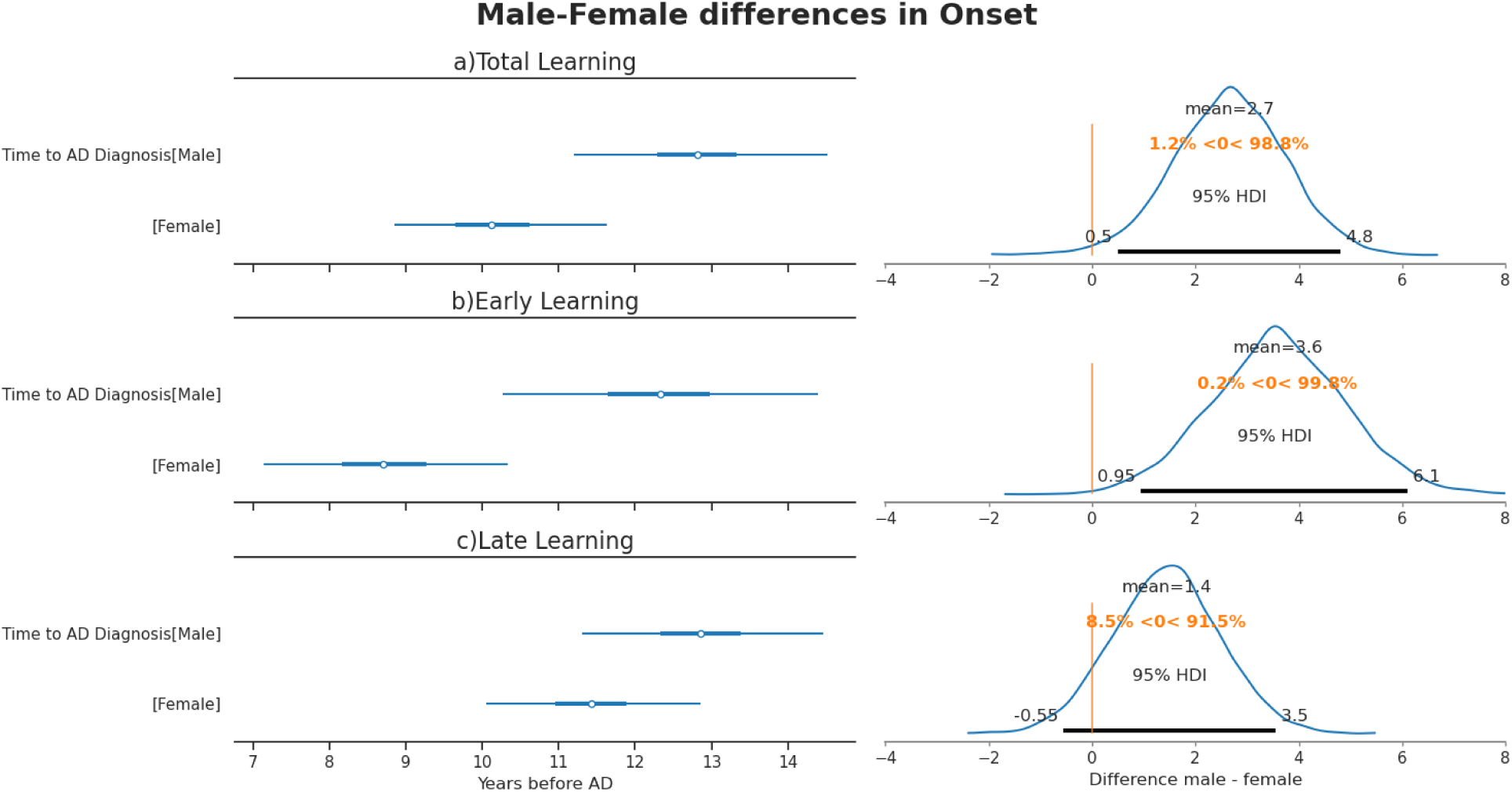
Posterior estimates of onset of verbal memory decline relative to AD diagnosis, by sex and RAVLT sub-scores. All models include Age, Education, and APOE ε4 as fixed effect covariates. Forest plots show the values of the posterior mean and 95% Highest Density Interval (HDI) for males, females, and their difference (male – female). Onset is expressed in years prior to AD diagnosis. Panel A shows Total Learning (sum of RAVLT trials 1-5), Panel B shows Early Learning (average of trials 1 and 2), and Panel C shows Late Learning (average of trials 4 and 5).

**Table 3:**
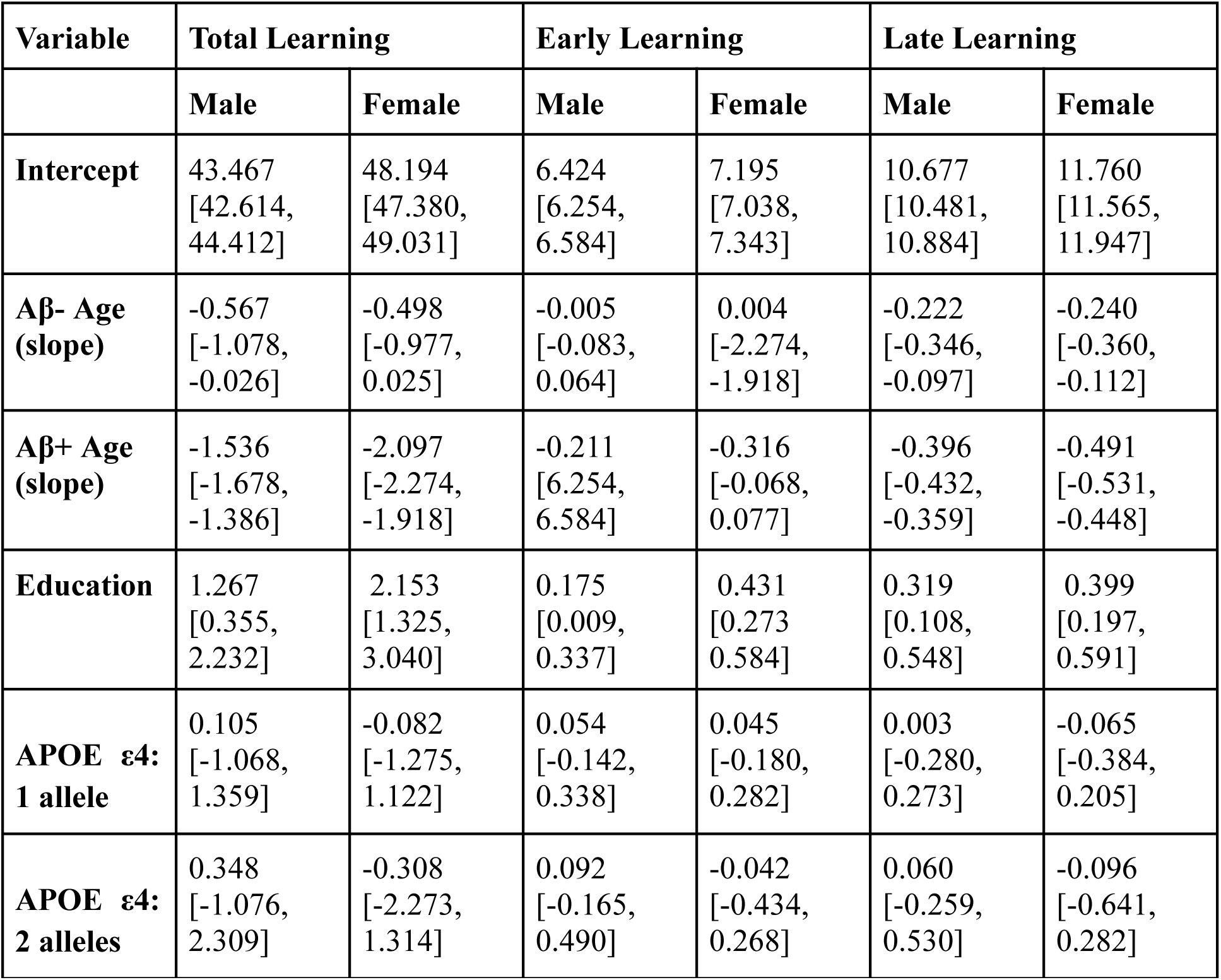
Sex-Stratified Model Estimates. ***Sex-stratified main effects coefficients of predictors of RAVLT subscores.*** Here the values represent the posterior mean with 95% High Density Intervals (HDI) in brackets. The model was fit using MCM sampling and fit was assessed with Leave-One-Out (LOO) validation.

### Females maintain normal verbal cognition significantly longer than males

Posterior estimates of onset time for Total Learning, Early Learning, and Late Learning decline controlling for age, education and APOE ε4 status are presented in Figure 2. Male-female differences of onset in Total and Early Learning decline were statistically significant. Females maintained normal performance 2.65 years longer in Total Learning (95% HDI = [0.5, 4.8], Fig. 2a) and 3.59 years longer in Early Learning (95% HDI = [0.95, 6.1], Fig. 2b). The male-female difference of 1.44 years for Late Learning (95% HDI = [−0.55, 3.55], Fig. 2c), was marginal with more than 5% of the posterior distribution in the negative range. Together, these results show that males begin to lose verbal cognition ∼13 years prior to an AD dementia diagnosis, while females will start to show symptoms ∼10 years before clinical detection.

### Females decline faster on all aspects of verbal learning

Posterior estimates of rate of decline for Total Learning, Early Learning and Late Learning controlling for age, education and APOE ε4 status are presented in Figure 3. For Total Learning (Fig. 3a), the posterior male-female mean difference was significant at 0.56 points per year (95% HDI = [0.34, 0.79], Table 3), representing a relative acceleration of 27–42% in females compared to males. On the Early Learning trials (Fig. 4b) the slope difference between males and females was 0.10 points per year (95% HDI = [0.06, 0.15], Table 3). Proportionally, this equates to a 47-52% accelerated post-onset working verbal memory decline in females compared to males. Late Learning (Fig. 4c) also indicated a significant 0.09 points per year difference (95% HDI = [0.04, 0.15], Table 3), corresponding to a 21-25% steeper slope in females relative to males. Females’ rapid decline in comparison to the males’ is suggestive of accelerated loss due to a depletion of cognitive reserve.

**Figure 3.**
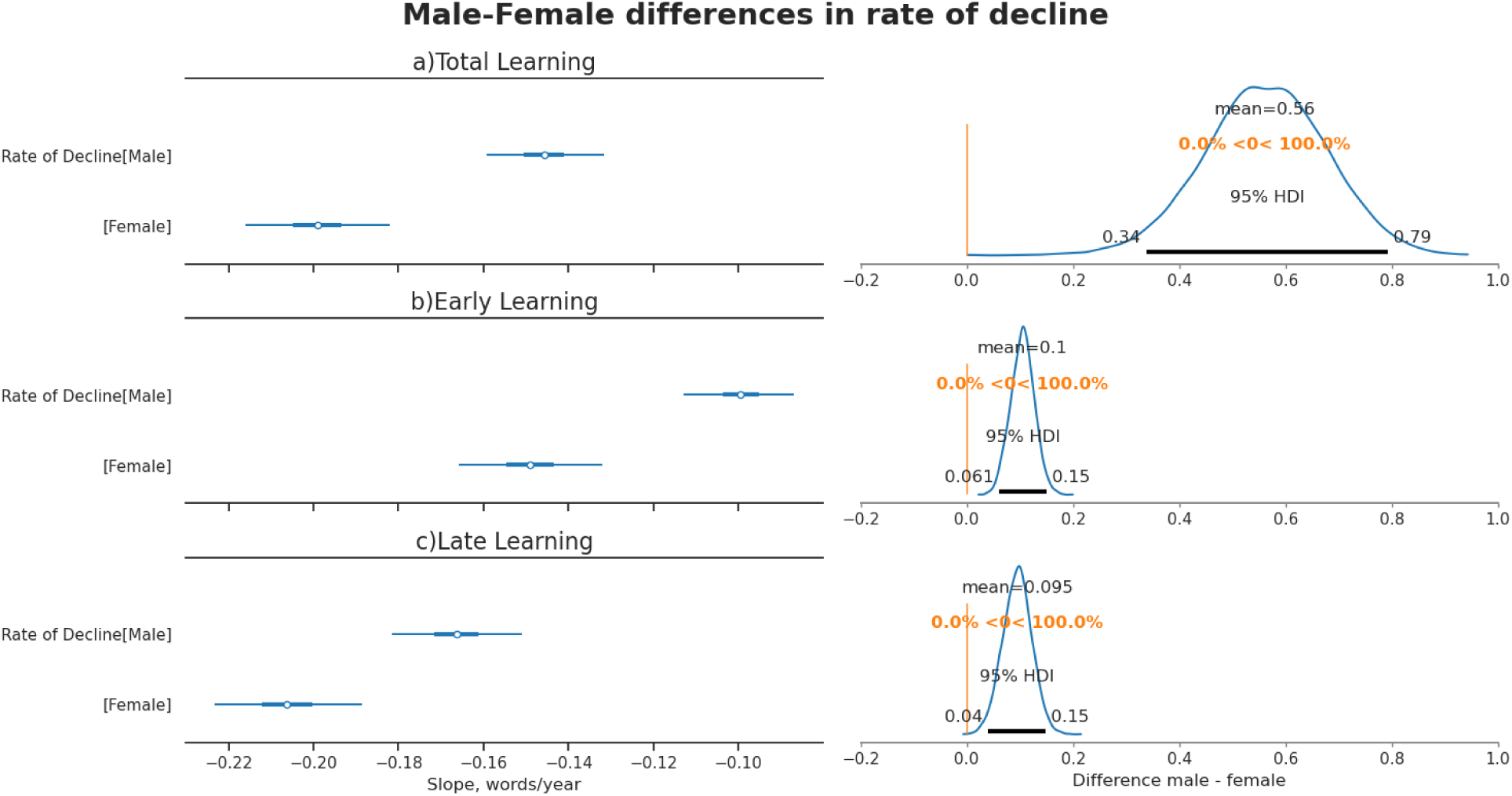
Posterior estimates of the rate of verbal memory decline, by sex and RAVLT subscore. All models include Age, Education, and APOE ε4 as fixed effect covariates. Forest plots show the values posterior mean and 95% Highest Density Interval (HDI) for males, females, and the difference (male – female). Decline is expressed in points per year. Panel A shows Total Learning (sum of RAVLT trials 1-5), Panel B shows Early Learning (average of trials 1 and 2), and Panel C shows Late Learning (average of trials 4 and 5).

## Discussion

The present study quantifies sex-specific trajectories of verbal memory decline in Aβ+ individuals. We demonstrated that (i) Aβ- females show a RAVLT total performance advantage (4.49/75 points, 6.0%, for RAVLT total), (ii) the onset of RAVLT total decline differs with females declining 2.65 years later than males, and finally, (iii) that once started, the slope of RAVLT total decline is steeper in females by 0.56 points ∼37% faster than males. These results align with emerging literature proposing that the female verbal memory advantage may serve as a form of dynamic cognitive reserve (Emrani & Sundermann, 2025). The delayed onset observed in females suggests that the protective effect of the verbal memory advantage is sufficient to delay detectable cognitive impairment measured by RAVLT. However, once these compensatory mechanisms are depleted, females suffer from a more precipitous rate of decline compared to males. This is evidenced further with the observed convergence of male and female RAVLT score at the official time of AD dementia diagnosis. This catch-up effect reveals cognitive reserve not as enduring protection, but as a longer fuse igniting faster neurodegeneration upon exhaustion.

In line with our secondary hypothesis, our results demonstrated larger effects in Early Learning in comparison to Late Learning (c.f, Fig 2). Thus, the apparent resilience could stem from Early Learning advantages related to executive strategies supporting working memory and early encoding that may obscure initial impairment and postpone clinical manifestation. It is of note that the effects of education, widely accepted to confer cognitive resilience, had a marginal impact on Early Learning in females. This pattern suggests that educational attainment earlier in life may be influential in building cognitive reserve in females with downstream effects later in life. Together, these results advance our understanding of the timing and cognitive mechanisms of the female verbal memory advantage and highlights the importance of sex-specific considerations in clinical assessment strategies. Our longitudinal analysis demonstrates the temporal dynamics of maintenance and subsequent disappearance of the verbal memory advantage, extending previous cross-sectional studies (Sundermann et al., 2016, 2017, 2019, 2020).

Our focus on immediate recall subscores of the RAVLT aligns with the perspective that encoding processes, rather than retrieval alone, play a critical role in episodic memory (Cherry et al., 2002; Lattmann-Grefe et al., 2024; Malek-Ahmadi et al., 2017; Pillai et al., 2014). Accomplishing the RAVLT depends not solely on episodic memory but relies heavily on working memory subprocesses to store new verbal memories (Putcha et al., 2019; Wolk et al., 2011). The fifteen-word list Early Learning trials (RAVLT 1-2) exceeds the phonological loop stores, probing strategic encoding to ensure RAVLT success. Thus, the mechanisms used to achieve better performance on the Early Learning RAVLT phase may allow participants to compensate for long-term storage deficits (Cherry et al., 2002; <u>Kljajevic et al. 2023</u>). Neuroimaging studies identified the temporal pole as a key structure for semantic organization necessary for effective performance on Early Learning, supporting robust long-term encoding (Putcha et al., 2019; Wolk et al., 2011). Conversely, Late Learning relies on the medial temporal lobe to consolidate information in long-term stores (Kwak et al., 2020). Our results mirror the progression of Braak staging, where early medial temporal lobe atrophy may be partially mitigated by the cognitive reserve in the temporal pole (Braak & Braak, 1991). The robust 3.59 years sex difference in Early Learning, compared to the marginal 1.44 year difference in Late Learning (Fig. 2), suggests that as medial temporal lobe dependent consolidation fails (Boona et al., 2011), females may utilize the semantic encoding strategies supported by the temporal pole (Jahn, 2013; Sundermann et al., 2016, 2017). The stark discrepancy in the rate of decline (∼50% vs ∼23%) implies that once this compensatory scaffold can no longer mask the underlying degeneration, the decline accelerates rapidly. Future work combining neuropsychological and neuroimaging approaches are necessary to determine whether sex differences in specific brain regions contribute to these behavioral patterns.

Sex differences in tau accumulation, a hallmark of AD pathology closely associated with cognitive decline, may help explain the differential trajectories of verbal memory decline (Lowe et al., 2013; Ossenkoppele et al., 2016; Yan et al., 2021). Evidence from an analysis of continuous global amyloid PET burden revealed higher early tau deposition in females independent of clinical diagnosis (Buckley et al., 2019; Ourry et al., 2025). Autopsy Braak staging findings also showed greater tau tangle burden independent of cognitive testing (Oveisgharan et al., 2018). These pathology driven approaches confirm that females exhibit greater tau burden at equivalent disease stages irrespective of diagnostic status. A previous study found that tau-RAVLT associations were stronger in Aβ+ MCI females, meaning that levels of tau accumulation better explain verbal memory decline in females than in males (Banks et al., 2021). Building on this, the same group investigated the relationship between residualized RAVLT and tau, demonstrating that females tolerate greater tau-PET burden before RAVLT decline (Digma et al., 2020). Taken together, these findings point to cognitive reserve despite accumulating pathology. However, our findings demonstrate that once AD dementia status is reached, male and female scores converge despite females starting at a higher baseline (Fig. 1). Indeed, it’s been reported that tau accumulates 10-15% faster in females (Coughlan et al., 2025), creating a paradoxical situation: later deviation from normal cognition but higher tau burden per RAVLT level. The accelerated accumulation of tau is in line with our results suggesting females experience a 25-50% steeper rate of cognitive decline relative to males (Fig. 3). Building on this mechanistic evidence, our longitudinal modeling approach precisely quantifies this female-specific accelerated decline across multiple facets of working verbal memory, providing robust temporal dynamics of the previously hypothesized female tipping point in AD progression (Nebel et al., 2018). This critical female point of accelerated change may reflect midlife biological transitions, such as menopause related estrogen loss which synergize with genetic risk to accelerate neuropathological burden (Merlini et al., 2025). Further research should aim to understand the neuropathological underpinnings of this steep decline, recognizing that neuropsychological assessments may lack sensitivity to detect AD symptoms in females within a treatable time window.

### Clinical Implications

Our results are in line with previous work that challenges the adequacy of current neuropsychological assessment practices (Emrani & Sundermann, 2025; Stricker et al., 2021; Sundermann et al., 2019). The temporal precision of our findings is not merely a statistical concern but represents a systematic bias that could deny women access to early interventions. For instance, in the Phase 3 clinical trial of Lecanemab, inclusion criteria required a classification of MCI or mild AD dementia as defined by non-sex-normalized assessments, including the WAIS Logical Memory test, which probes working and episodic verbal memory (van Dyck et al., 2023). Prior work has suggested that the rapid rate of cognitive decline may reflect higher underlying neuropathology and our results confirmed that sex-blind assessments may classify women as MCI only once they are further along in the disease process (Banks et al., 2021; Digma et al., 2020; Sundermann et al., 2019). This may explain why Lecanemab appears to benefit males more than females (Andrews et al., 2025). Our findings demonstrate that sex-blind verbal memory assessments with RAVLT may be insufficient for detecting subtle change in early AD, particularly in females. As precision medicine advances, diagnostic tools must account for sex-specific differences in cognitive reserve to avoid systematic underdiagnosis of women in the earliest, most treatable stages of disease. It is imperative that clinicians integrate the sex-specific considerations highlighted here into their assessments to prevent delayed diagnosis and treatment in women. Consistent with initiatives by the National Institute in Aging (Jack et al., 2018), we also advocate for the use of targeted biomarkers of AD rather than relying solely on cognitive measures.

### Study Limitations

While this study addresses critical gaps in the literature by modelling longitudinal change in verbal memory across sexes, it does not provide evidence for the neural basis of such differences. Future studies should test the theories proposed above by investigating tau pathology in relation to cognitive decline measured with list learning assessments. The nature of combining two cohorts did not allow us to do this because there currently does not exist a harmonized measure for Tau accumulation (Leuzy et al., 2024). Additionally, the cohorts we used were made primarily of educated Caucasians, which may reduce generalizability. Finally, we did not include RAVLT Delayed Recall in our analyses, despite its established importance in assessing cognitive decline in AD, because the data exhibited a floor effect, with a substantial number of zeros.

## Conclusion

This study provides a comprehensive longitudinal characterization of sex differences in verbal memory decline trajectories relative to AD onset. Our findings support the hypothesis that women’s superior verbal memory abilities serve as a form of cognitive reserve that delays clinical detection of AD-related decline but results in steeper deterioration once compensatory mechanisms are overwhelmed. These sex-specific patterns have significant implications for clinical practice and highlight the need for sex-aware approaches to AD assessment and intervention.

## Data Availability

All data describe in the manuscript are open access. The code will also be made publicly available.

## Acknowledgements

The present study was supported through the Canadian Institutes of Health Research funding agency and donations from Louise et Andre Charron family foundation.

## Appendix

**Equation A1. Hierarchical Bayesian model predicting RAVLT performance**

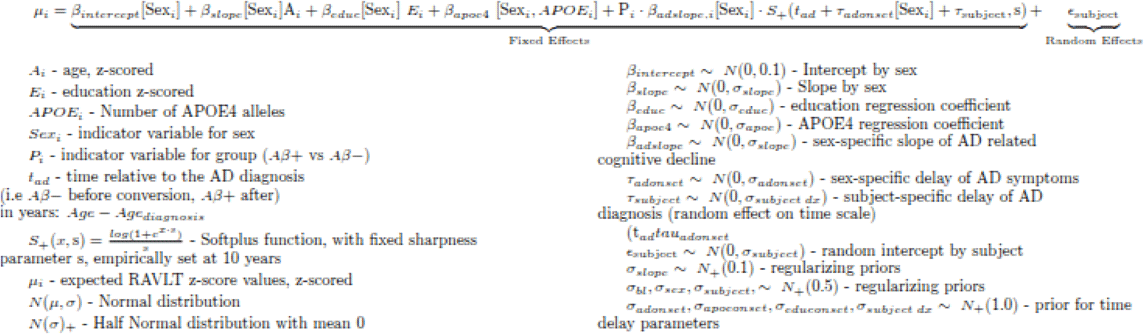

***Equation A1: In depth hierarchical Bayesian model specification.*** This model predicts scores for RAVLT Total, Early and Late Learning as a function of Age, Education, APOE status and group (Aβ- vs Aβ+). Analysis code is publicly available at this link: https://github.com/NIST-MNI/ravlt_ad_trajectory. Data used in the preparation is available upon request from ADNI and PREVENT-AD.

**Figure A1:**
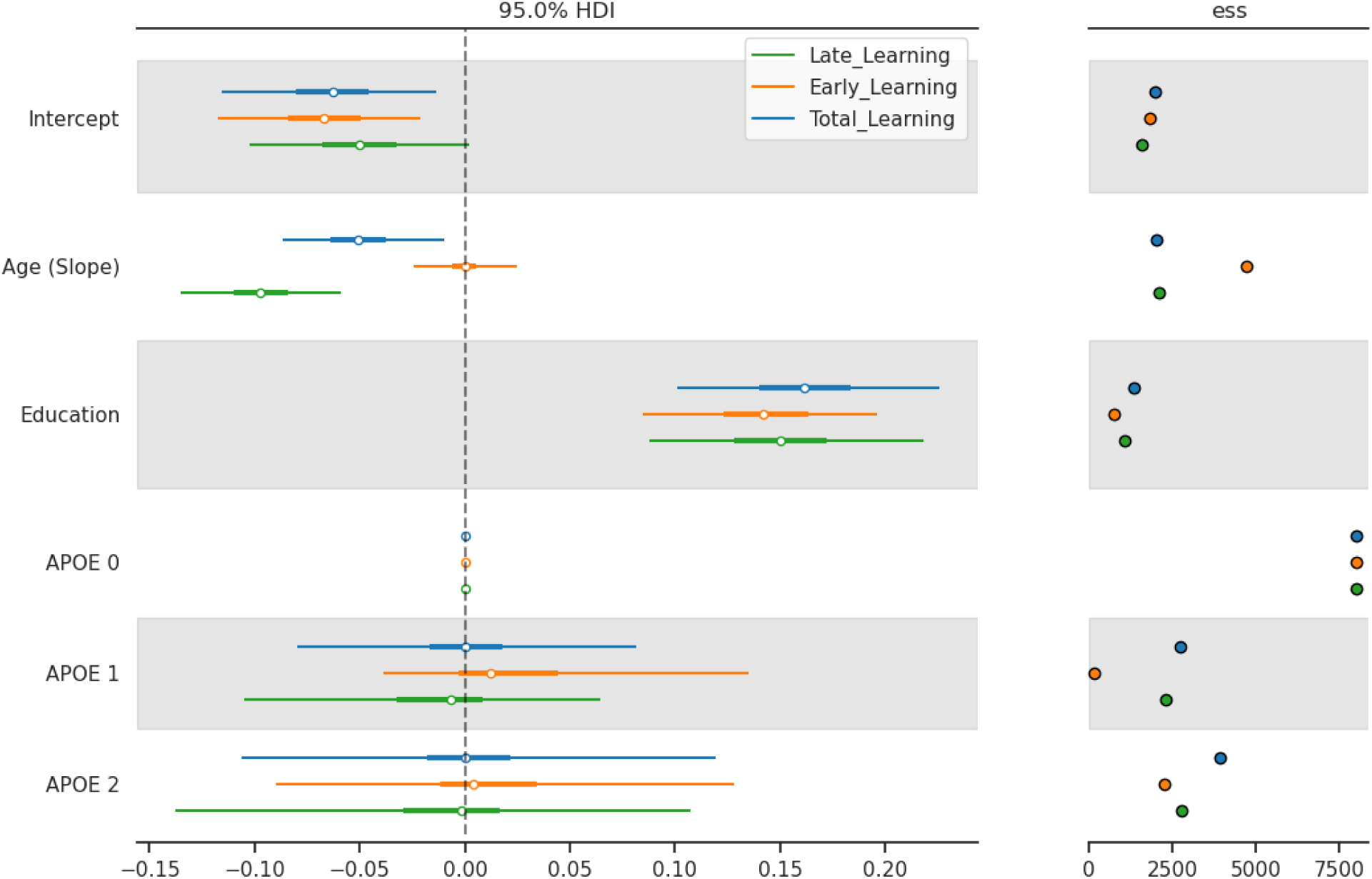
Forest Plot of Z-scored General Model Estimates. ***Posterior distribution of global model coefficients for RAVLT subscores.*** Points represent the posterior mean and horizontal bars indicate 95% High Density Interval (HDI). A vertical dashed line at zero represents the null effect threshold. Intervals excluding this line are considered to significantly impact RAVLT performance. Model convergence was verified via Effective Sample Size (ESS) diagnostics.

**Figure A2:**
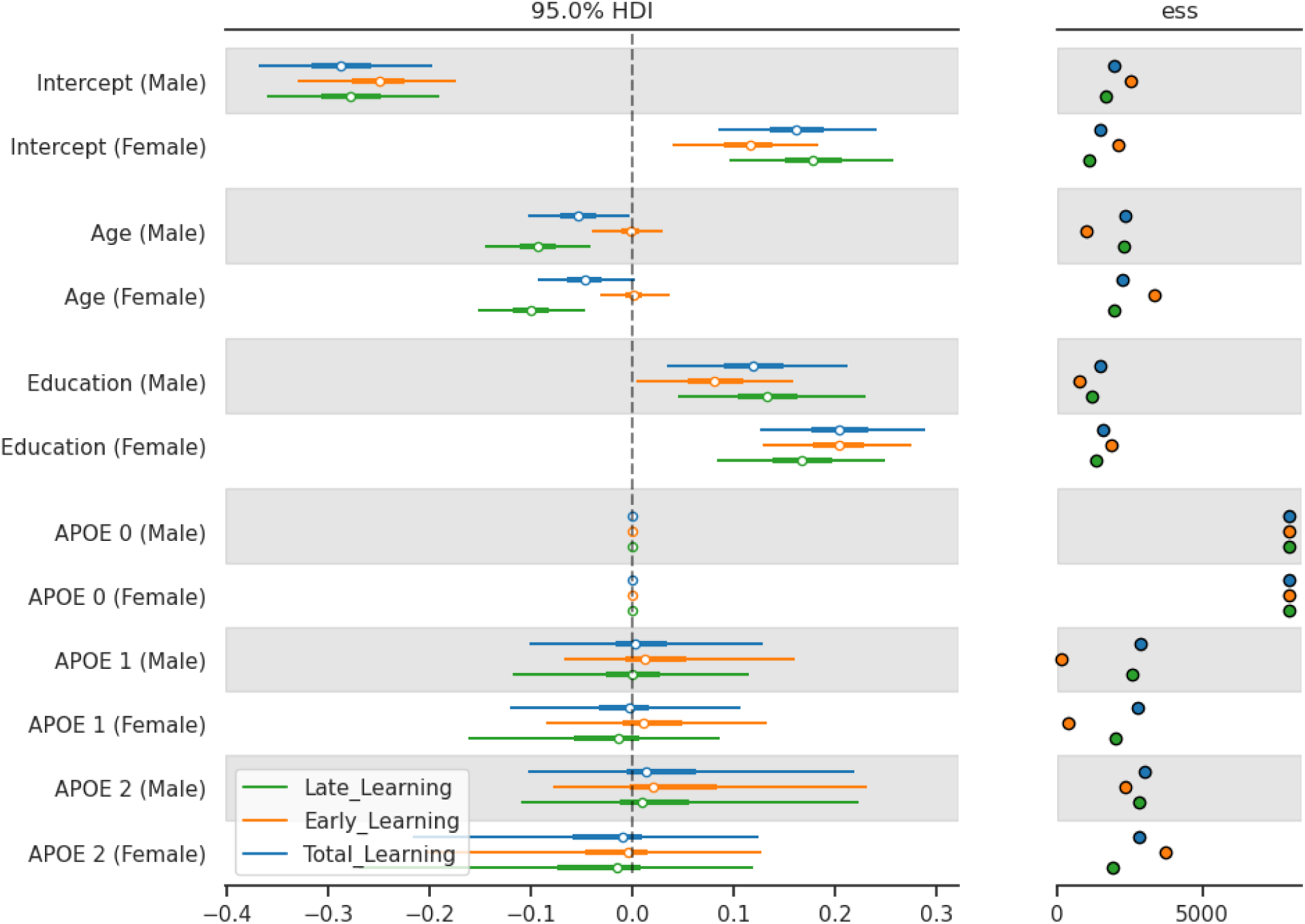
Forest Plot of Z-scored Sex-stratified model estimates. ***Posterior distribution of sex-stratified model coefficients for RAVLT subscores.*** Points represent the posterior mean and horizontal bars indicate 95% High Density Interval (HDI). A vertical dashed line at zero represents the null effect threshold. Intervals excluding this line are considered to significantly impact RAVLT performance. Model convergence was verified via Effective Sample Size (ESS) diagnostics.

