## Supplemental figures for "Sex specific trajectories of verbal memory decline suggest masking of Alzheimer’s Disease symptoms in females" (from another version of this preprint: version 1, 2025-11-09)

**Supplementary material**

1. **eMethods. Hierarchical Mixed-Effects Model for Verbal Memory Decline**
2. **eFigure 1. Posterior male-female difference estimates for all parameters**
3. **eFigure 2. Forest plots showing the effect of APOE ε4 status on model divergence from cognitively normal trajectories.**

**eMethod. Hierarchical Bayesian Mixed-Effects Model for Verbal Memory Decline**

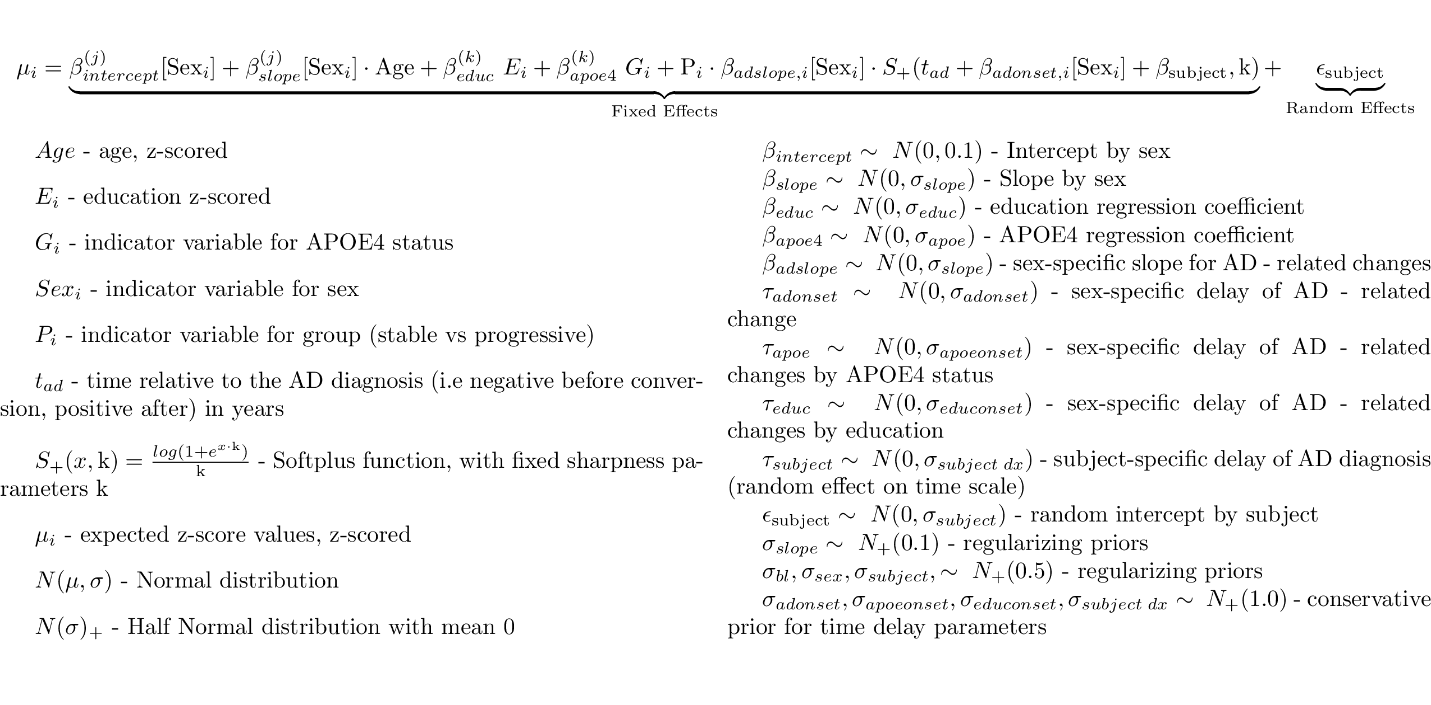

**eMethod*. Hierarchical Bayesian Mixed-Effects Model for Verbal Memory Decline.*** A probabilistic framework integrating demographic and genetic covariates with nonlinear time dynamics demographic and genetic covariates with nonlinear time dynamics to estimate subject-specific cognitive decline. The model leverages SoftPlus transformations and regularizing priors or robust inference.

**eFigure 1. Posterior male-female difference estimates for all parameters**

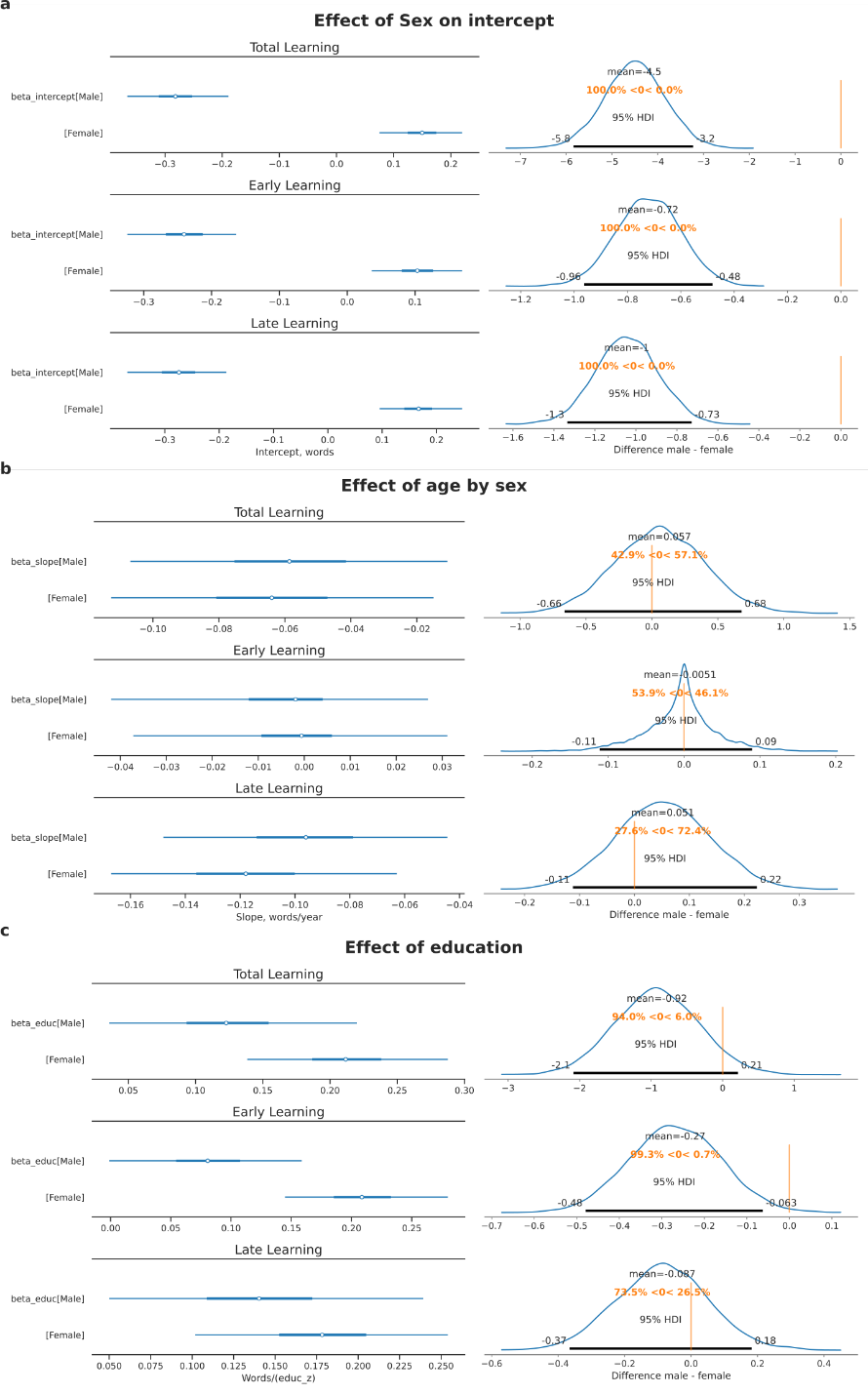

***eFigure 1.*** ***Posterior male-female difference estimates for all parameters.*** Panel A shows the differential male-female effect in sex at baseline. Panel B shows the differential male-female effect in age/time. Panel C shows the differential male-female effect of education. Panel D shows the differential male-female effect of APOE genotype. Black bars within the distribution represent the 95% HDI.

**eFigure 2. Forest plots showing the effect of APOE ε4 status on model divergence from cognitively normal trajectories.**

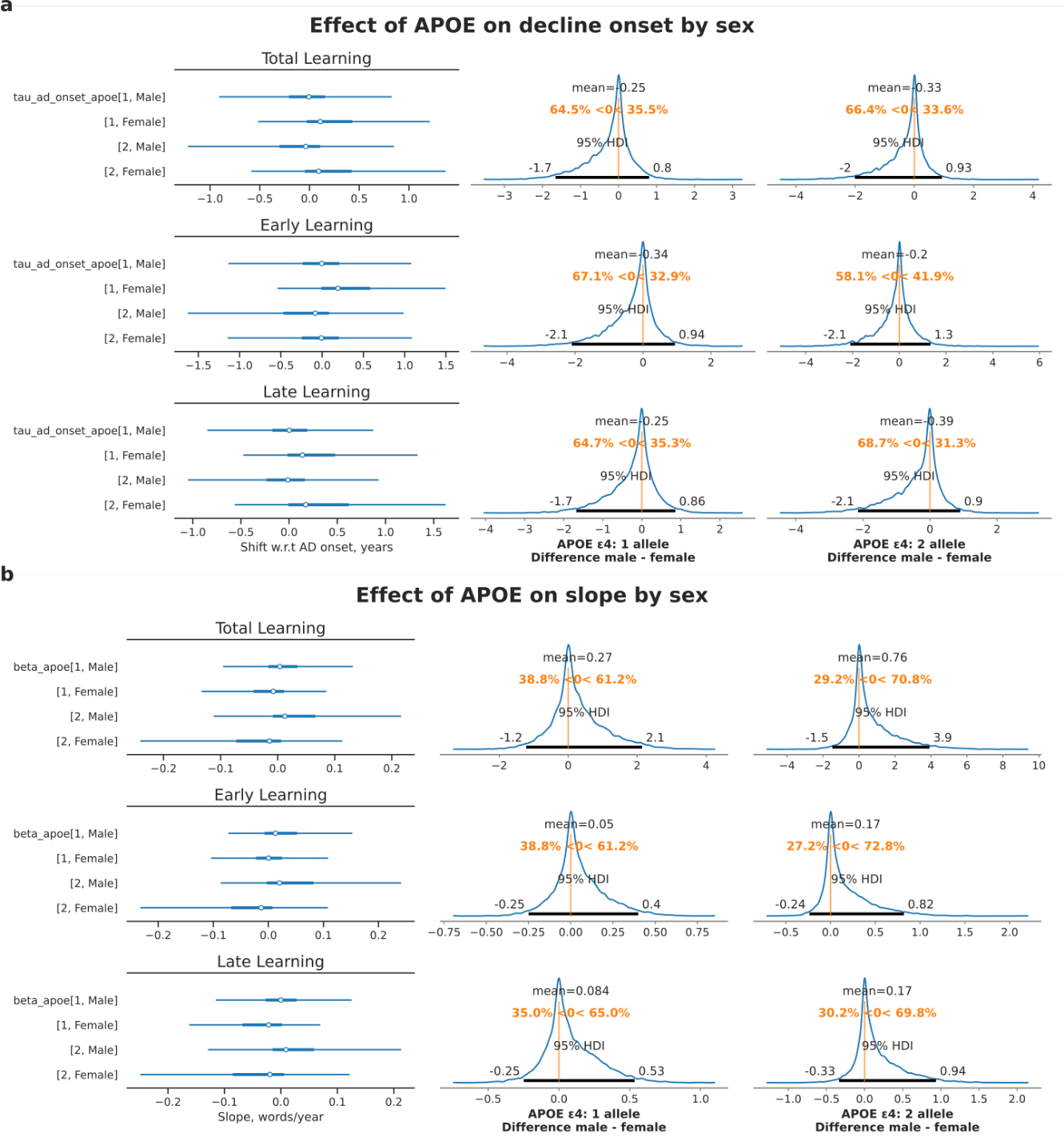

***eFigure 2. Forest plots showing the effect of APOE ε4 status on model divergence from cognitively normal trajectories.*** Panel A shows the male-female difference in the effect of APOE **ε4** on the timing of decline. Panel B shows the male-female difference in the effect of APOE **ε4** on the slope. The black bars inside the distributions represent the 95% HDI.
